# The effects of the infarct volume on cytokines and immune status in patients with acute ischemic stroke

**DOI:** 10.1101/2023.10.16.23297078

**Authors:** Xingqi Su, Lingmin Zhao, Di Ma, Jiulin You, LiangShu Feng, Jing Wang, YuLei Hao, XinYu Wang, JiaChun Feng

## Abstract

**Objective:** To investigate and analyze the effects of the infarct volume on cytokines and immune status in patients with acute ischemic stroke.

**Methods:** Patients with acute ischemic stroke that presented within 72 h of onset from October 2017 to October 2019 were enrolled. Patients with severe cerebral infarction (large-area cerebral infarction) (n=34) were enrolled and categorized as group A; Additionally, 33 patients with non-large-area cerebral infarction with matching baseline characteristics (sex and age) to group A were included in group B. We measured IL-2, IL-4, IL-6, IL-10, IL-17A, TNF-α, and IFN-γ levels in serum using a cytometric bead array.In addition,we compared the absolute value of lymphocytes (LYM#), lymphocyte percentage (LYM%), neutrophil/lymphocyte ratio (NLR), cytokine levels, and immune status indicators (IFN-γ IL-4 ratio, TNF-α/IL-4 ratio, and TNF-α/IL-10 ratio) between groups A and B, and evaluated the effect of infarct size on inflammatory factors and immune status.

**Results:** Compared with group B, the LYM# and LYM% in group A were significantly lower, and the NLR and cytokines (IL-2, IL-4, IL-6, IL-10, IL-17A, TNF-α, and IFN-γ) levels were significantly higher. TNF-α/IL-4 ratio was significantly lower, and the IFN-γ/IL-4 ratio (P=0.09) and TNF-α/IL-10 ratio (P=0.146) in group A demonstrated a decreasing trend although not significant.

**Conclusions:** The immune status of patients with acute cerebral infarction is related to the infarct volume; patients with large-area cerebral infarction are more likely to develop immunosuppression.

## 1. Introduction

According to the all-cause mortality statistics of the World Health Organization (WHO), stroke is the second leading cause of death globally, accounting for 87%[1], and its high disability rate significantly increases the social burden. Because immunity is crucial in cerebral infarction and, in turn, is regulated by it, the immune mechanism has become a target that may benefit clinical treatment and has attracted increasing interest from researchers.

Notably, immunity is crucial in the occurrence, progression, and outcome of ischemic stroke. The basic pathology of atherosclerotic cerebral infarction is atherosclerosis. Various immune cells, including lymphocytes and mononuclear macrophages, are involved in the formation of atherosclerotic plaques and the rupture of unstable plaques[2,3], an important cause of atherosclerotic cerebral infarction[4]. Immunity contributes to the pathogenesis of cerebral infarction and affects prognosis[5–8]. After local cerebral infarction, astrocytes and microglia, which are involved in the release of cytokines and chemokines[9], are rapidly activated, attract immune cells from the peripheral circulation to the ischemic area, and participate in inflammatory damage to the brain tissue[10]. In addition, in the recovery from chronic cerebral infarction, the immune status affects the brain tissue microenvironment, regulates neurogenesis[11,12] and angiogenesis[13], and affects the autoimmune response to brain tissue antigens after stroke, mediating the long-term prognosis[5,6]. Notably, as a stress event, ischemic stroke regulates systemic immunity through the sympathetic nervous system and the hypothalamus-pituitary-adrenal axis, which is an important mechanism of post-stroke immunosuppression, leading to increase mortality of patients with acute cerebral infarction [7,8,16-19]. We know that the size of different infarcts also affects the treatment and prognosis of acute ischemic stroke,(PMED: 28665171)In this study, we investigate and analyze the effects of the infarct volume on cytokines and immune status in patients with acute ischemic stroke.This enables the clinicians to make informed decisions in selecting immunotherapy approaches and tailor rehabilitation strategies to optimize patient outcomes by considering different infarct sizes.

## 2. Materials and methods

### 2.1 Research objects and samples

#### Inclusion criteria

1. Patients with acute ischemic stroke presenting with an onset time of ≤72 h in the Department of Neurology of the First Hospital of Jilin University(Changchun,China) from October 2017 to October 2019.
2. Patients who voluntarily participated in the study.
3. Patients aged 18–80 years old, regardless of sex.

#### Exclusion criteria

1. Intracranial artery dissection, aneurysm, vascular inflammatory diseases, vascular malformations, and other neurological diseases.
2. Patients who have undergone thrombolysis or thrombectomy.
3. Patients with or suspected cerebral embolism.
4. Abnormal liver and kidney function; AST or ALT 3 times higher than the upper limit of normal value; creatinine clearance rate of <0.6 mL/s; blood creatinine level >265 μmol/L.
5. Patients with autoimmune diseases, who have received immunotherapy, or with malignant tumors.
6. Patients with severe trauma or have undergone major surgery recently.

#### Sample collection

Peripheral blood (2–3 mL) of the patients were collected immediately after admission using a coagulation blood collection tube. After centrifugation (4 °C, 3000 r/min, 5 min), the supernatant (serum samples) was collected and cryopreserved at - 80 °C.

### 2.2 Reagents and equipment

BD FACS Calibur flow cytometer (BD Company, USA); low-temperature high-speed centrifuge (Heraeus Sepaeeh, Germany); low-temperature refrigerator (Haier Company, Qingdao, China); BD™ Cytometric Bead Array (CBA) human Th1/Th2/Th17 kit (BD Company, USA); BD™ Cytometric Bead Array (CBA) Human Soluble Protein Master Buffer kit, (BD Company, USA).

### 2.3 Determination of cytokines in serum samples by CBA method

The BD™ Cytometric Bead Array (CBA) human Th1/Th2/Th17 kit was used to determine the cytokine (IL-2, IL-6, IL-4, IL-10, IL-17A, IFN-γ, TNF-α) levels in each serum sample according to the manufacturer’s instructions.

### 2.4 Experimental group

A total of 67 patients presenting with an onset time of 72 h were included in this study;patients with large-area cerebral infarction were included in the large-area cerebral infarction group and categorized as group A. A total of 34 patients, including patients with anterior circulation infarction and posterior circulation infarction, were included. For anterior circulation infarction, the maximum diameter of the infarct was >3 cm (25 participants), and regarding posterior circulation infarction, the maximum infarction cross-section was >1/3 of the brainstem (9 participants).

Additionally, 33 patients with non-large-area cerebral infarction with matching sex and age with Group A were included in the non-large-area cerebral infarction group and categorized as Group B, including patients with anterior circulation infarction and posterior circulation infarction. For anterior circulation infarctions, the largest infarct diameter was <3 cm (27 participants), and for posterior circulation infarctions, the largest brainstem infarct area was <1/3 of the cross-sectional area (6 participants).

### 2.5 Statistical analysis and graph

Statistical analysis was performed using IBM SPSS Statistics version 25.0 (IBM Corp., Armonk, N.Y., USA), and GraphPad Prism version 7 (GraphPad Software, Inc., La Jolla, CA, USA) was used for graphing. The counted data were presented as the number of cases (n) and percentages (%). Measurement data with normal distribution are presented as 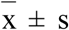 and measurement data that are not normally distributed are presented as the median and quartile M (Q25–Q75). For counting data, the Chi-square test (κ2 test) was used to compare groups. For the comparison between the two groups of measurement data, the t-test was used for normally distributed data, and the nonparametric rank-sum test (Mann–Whitney U test) was used for non-normal distribution. For the comparison between the three groups of measurement data, a one-way analysis of variance (ANOVA) was used for the normal distribution, and the variance was uniform. The LSD-t test was used for the pairwise comparison of the mean between the groups.

The groups showed statistically significant differences, with a P-value of <0.05 indicating statistical significance; for pairwise comparison of the mean between the groups, the statistical difference was based on the unadjusted P-value <0.017 and the adjusted P-value <0.05.

## 3. Results

We compared the expression levels of various cytokines, including IL-2, IL-4, IL-6, IL-10, and IL17A, in the large-area infarction group (group A) and the non-large-area infarction group (group B) based on the same baseline level. Furthermore, we compared the absolute lymphocyte value (LYM#), lymphocyte ratio lymphocyte percentage (LYM%), neutrophil/lymphocyte ratio (NLR), IFN-γ/IL-4 ratio, TNF-α/IL-4 ratio, and TNF-α/IL-10 ratio between groups A and B.

The results indicated the following: (1) The cytokine levels in group A were significantly higher than in group B, as shown in Table 2 and Fig 1. These differences were considered statistically significant; P< 0.05. (2) Group A exhibited significantly lower LYM# and LYM% than group B, and the NLR was significantly higher. These findings are presented in Table 3 and Fig 2 and showed statistically significant differences. (3) The TNF-α/IL-4 ratio in group A was significantly lower than in group B, which showed a statistically significant difference. The TNF-α/IL-10 ratio and IFN-γ/IL-4 ratio in group A demonstrated a decreasing trend but showed no statistically significant differences (Table 4 and Fig 3).

**Table 1.** Baseline data between patients in group A and group B.

| Baseline | Group A | Group B | P-value |
| --- | --- | --- | --- |
| Sex, Male (n, %) | 19 (55.88%) | 24 (72.7%) | 0.151 |
| Female (n, %) | 15 (44.12%) | 9 (27.3%) |  |
| Hypertension (n, %) | 23 (67.65%) | 26 (78.79%) | 0.304 |
| Diabetes (n, %) | 10 (29.42%) | 10 (30.30%) | 0.936 |
| Smoking history (n, %) | 14 (41.17%) | 13 (39.39%) | 0.882 |
| Age (years) | 63.5 (57–69) | 62.5 (56.25–69) | 0.768 |
| LDL-C (mmol/L) | 3.06 (2.72–4.34) | 2.9 (2.29–3.86) | 0.075 |
| Hcy (μmol/L) | 14.45 (10.93–19.65) | 15.3 (13.4–20.8) | 0.166 |
| Fasting blood glucose (mmol/L) | 5.84 (5.23–7.93) | 5.71 (4.96–6.61) | 0.349 |
| NIHSS score | 13 (11–16.25) | 4 (3–8) | 0.000 |

**Table 2.** Serum cytokine levels of patients of group A and group B.

| Cytokine level | Group A | Group B | U-value | P-value |
| --- | --- | --- | --- | --- |
| **IL-6 (pg/mL)** | 32.35 (9.69–56.60) | 9.81 (3.09–28.60) | 320.00 | 0.003 |
| **IL-4 (pg/mL)** | 4.02 (3.45–4.76) | 0.58 (0.31–0.80) | 27.00 | 0.000 |
| **IL-2 (pg/mL)** | 1.96 (1.75–2.19) | 0.55 (0.32–0.98) | 84.50 | 0.000 |
| **IL-17A (pg/mL)** | 8.15 (6.30–9.66) | 3.25 (1.37–5.08) | 142.00 | 0.000 |
| **IFN-γ (pg/mL)** | 2.58 (2.30–2.99) | 0.61 (0.22–0.95) | 13.00 | 0.000 |
| **TNFα (pg/mL)** | 2.63 (2.23–3.15) | 1.71 (0.80–2.82) | 284.00 | 0.001 |
| **IL-10 (pg/mL)** | 3.43 (2.66–4.52) | 1.31 (0.89–2.46) | 169.00 | 0.000 |

Mann–Whitney U test was used. The cytokine levels in group A were significantly higher than in group B.

**Table 3.** LYM#, LYM%, and NLR between group A and group B.

| Indicators | Group A | Group B | U-value | P-value |
| --- | --- | --- | --- | --- |
| **LYM# ($\times 10^9$)** | 1.41 (1.05–1.79) | 1.76 (1.43–2.35) | 324.50 | 0.003 |
| **LYM%** | 0.14 (0.08–0.22) | 0.26 (0.21–0.33) | 214.00 | 0.000 |
| **NLR** | 5.27 (3.26–10.79) | 2.61 (1.73–3.36) | 204.00 | 0.000 |

Mann–Whitney U test was used. The LYM# and LYM% of the large-area group (group A) were significantly lower than those of the non-large-area group (group B), while the NLR of group A was significantly higher than that of group B.

**Table 4.** Indicators of immune status of patients in group A and group B.

| Indicators | Group A | Group B | U-value | P-value |
| --- | --- | --- | --- | --- |
| **TNF-α/IL-4** | 0.69 (0.54–0.83) | 2.50 (1.62–5.99) | 137.00 | 0.000 |
| **TNF-α/IL-10** | 0.80 (0.53–1.12) | 1.05 (0.53–3.06) | 445.00 | 0.146 |
| **IFN-γ/IL-4** | 0.66 (0.52–0.82) | 1.07 (0.40–2.06) | 426.00 | 0.090 |

Mann–Whitney U test was used. The TNF-α /IL-4 ratio in group A was significantly lower than in group B; the TNF-α /IL-10 and IFN-γ /IL-4 ratios in group A showed a downward trend.

**Fig 1.**
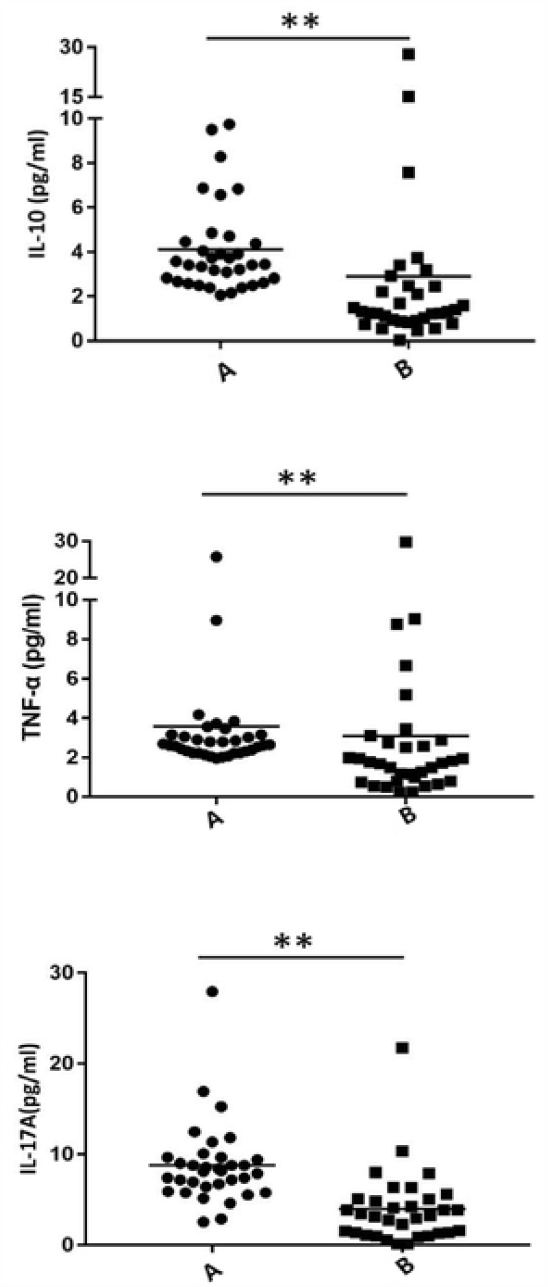
Serum cytokine levels of groups A and group B patients. ** P<0.01.

**Fig 2.**
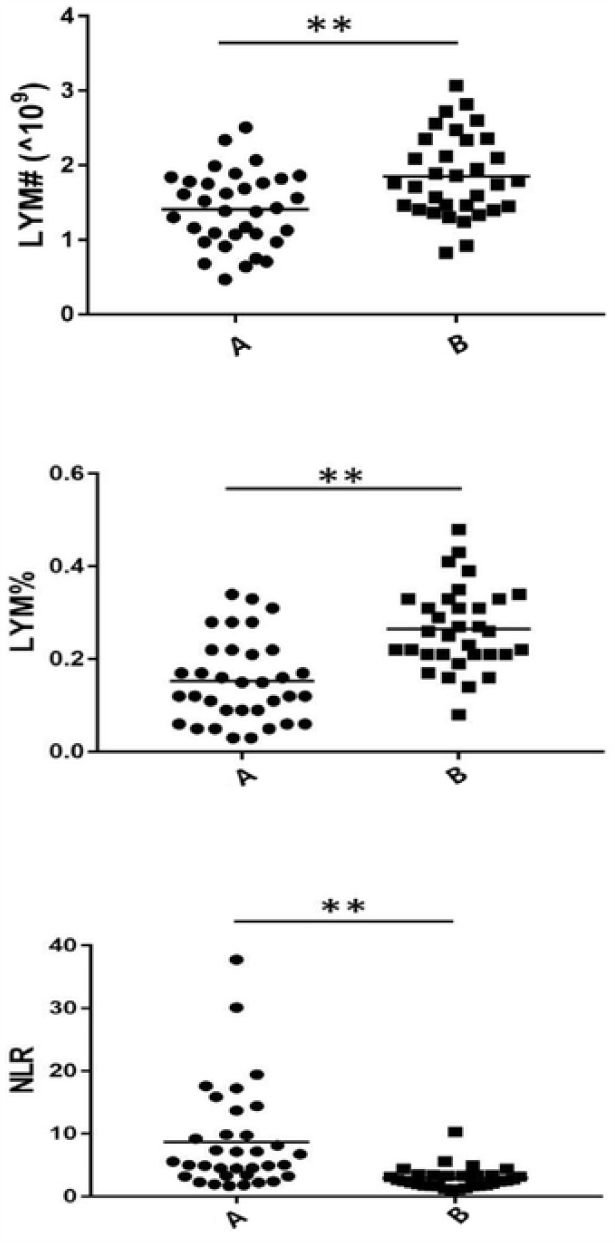
LYM#, LYM%, NLR of group A and group B. ** P<0.01

**Fig 3.**
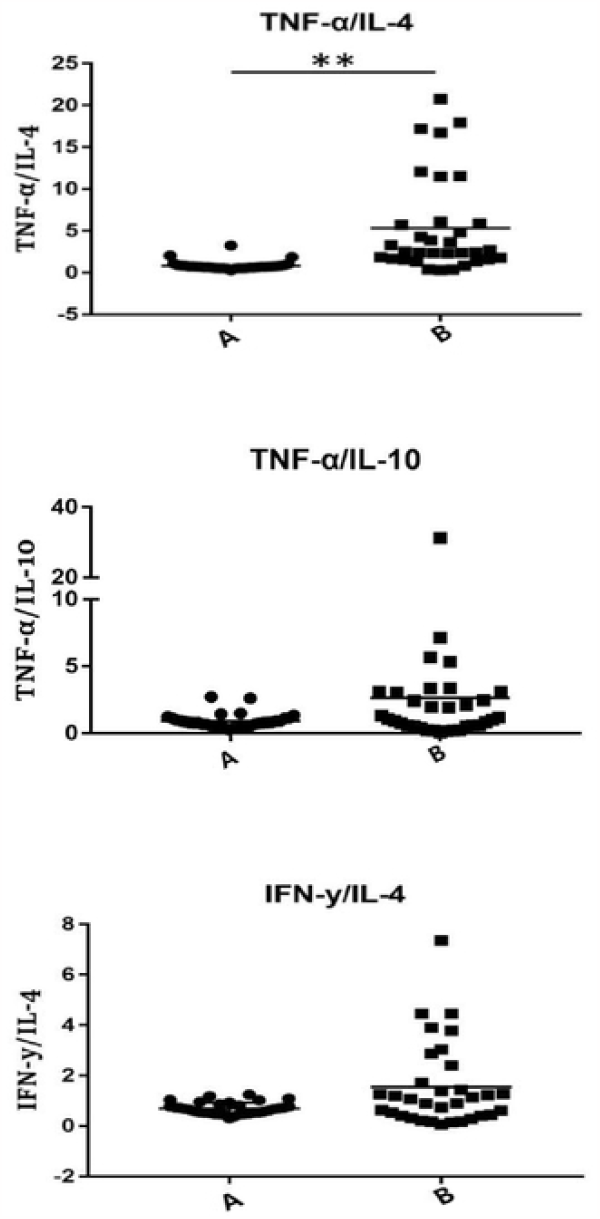
Immune status of patients in group A and group B. ** P<0.01.

## 4. Discussion

Ischemic stroke is a common cerebrovascular disease; it causes irreversible brain tissue damage. The complex pathological mechanism includes excitatory glutamate toxicity, calcium overload, oxidative stress, and inflammatory damage.

Inflammatory reaction is crucial in brain tissue injury following infarction. The immune state regulates local pathophysiological changes and significantly affects the outcome of ischemic tissues. The proinflammatory state aggravates local injury; however, the anti-inflammatory state protects the local brain tissue, promotes the removal of necrotic cells, and prevents further damage. Inflammation of the central nervous system and peripheral systemic immunity are closely linked and reflect the delicate and ingenious integrity of the human body. For example, the blood-brain barrier (BBB) is compromised after cerebral infarction and peripheral immune cells invade the brain tissue. The Th cells that invade the central nervous system secrete several cytokines and interact with microglia to initiate microglial polarization [14,15]; this affects the degree and outcome of the local brain tissue damage.

The relationship between the nervous and immune systems is such that the immune system affects the pathophysiological changes of cerebral infarction and is, in turn, affected by cerebral infarction events, such as immunosuppression after stroke. The underlying mechanism is not fully understood.Post-stroke immunosuppression is a self-protective mechanism for suppressing central inflammation; however, it alters peripheral immune status and further affects the prognosis in patients with acute cerebral infarction[7–8].

Considering the role of immune factors in the course of cerebral infarction, investigating the immune mechanisms has increasingly attracted the interest of researchers. The body’s immunity is affected by several factors, and some related factors can be controlled artificially; therefore, the immune mechanism may be a potential clinical treatment target. In this study, we evaluated the levels of various cytokines, including IL-2, IL-6, TNF-α, IFN-γ, IL-4, IL-10, and IL-17A in patients presenting with acute cerebral infarction within 72 h of onset, and calculated the INF-γ/IL-4, TNF-α/IL-4, and TNF-α/IL-10 ratios. The IFN-γ/IL-4 ratio indicates the balance between the subpopulations of helper T cells, reflecting the relative function between the Th1 and Th2 subpopulations. IFN-γ is an essential proinflammatory cytokine secreted by Th1-type cells, while IL-4 is a relatively specific cytokine of Th2-type cells and exerts an anti-inflammatory effect. The initial Th cells, Th0 cells, are regulated by several factors and differentiated into subpopulations. The most significant factor is the type and level of cytokines and the balance between each cytokine[20]. Therefore, the IFN-γ/IL-4 ratio is of great significance. Furthermore, the mononuclear macrophage system is divided into M1- and M2-type responses and is regulated by the Th1/Th2 immune status. Proinflammatory M1-type macrophages are polarized by binding to lipopolysaccharide (LPS) or Th1-type cytokines (such as IFN-γ), producing proinflammatory cytokines including IL-1β, IL-23, TNF-α; anti-inflammatory M2 macrophages are polarized by Th2-related cytokines, such as IL-4 and IL-13, and secrete anti-inflammatory cytokines such as IL-10 and TGF-β[21], promoting tissue healing and inflammation resolution[22]. Therefore, the M1/M2 type immune response can be evaluated by the TNF-α/IL-4 ratio and the TNF-α/IL-10 ratio.

In this study, by collecting laboratory test results, imaging data, and medical history, we discovered that the levels of various inflammatory factors and immune status in patients with acute cerebral infarction were affected by the infarct size.

The infarct size significantly affects the immune system. While several studies have focused on the effect of infarct size on adaptive immune function impairment, we explored its effects on the immune status and the balance between proinflammatory and anti-inflammatory responses. Fluctuations in various cytokine levels in the peripheral circulation can reflect changes in peripheral immunity following stroke. Our results showed that the levels of cytokines (IFN-γ, TNF-α, IL-2, IL-4, IL-6, IL10, IL-17A) were significantly higher in the group with large-area cerebral infarction than those in non-large-area cerebral infarction group; however, the extent of increase varied for each cytokine. Furthermore, compared to the baseline-matched non-large cerebral infarction group, the absolute value and percentage of lymphocytes decreased significantly in patients with large cerebral infarctions; however, the NLR was significantly higher in the large-area cerebral infarction group. Neutrophils are crucial in innate immunity, and the NLR reflects the degree of immune disorder. A higher NLR indicated a greater degree of adaptive immunity impairment. Our findings suggest that patients with large-area cerebral infarctions have more impaired adaptive immune functions, and the relative levels of cytokines affect the immune status (adaptive immunity). Additionally, we calculated the IFN-γ/IL-4, TNF-α/IL-4, and TNF-α/IL-10 ratios. The data showed that the TNF-α/IL-4 ratio was significantly decreased in the group with large-area cerebral infarction, indicating a more anti-inflammatory balance. Notably, the IFN-γ/IL-4 and TNF-α/IL-10 ratios were decreased, suggesting a shift towards an anti-inflammatory response in patients with severe infarction.

The immune response is involved throughout cerebral infarction and plays an important role in the process of disease transformation with a great degree of variation. Therefore, it is a potential target for improving the prognosis and outcomes of neurological function. However, at present, there is no systematic research on the effects of the infarct volume on cytokines in patients with acute ischemic stroke.and there is a lack of a deeper understanding of the impact of the immune status on clinical adverse events. Cerebral infarction is a complex disease; several conditions induce it, and the pathological mechanism after its onset is complicated.

This study collected first-hand clinical information on patients and systematically analyzed that severity of infarction may affect the serum cytokine levels and immune status of patients with acute cerebral infarction, and explored how it regulates immunity. This study provides valuable clinical data for related fields and enables the clinicians to make informed decisions in selecting immunotherapy approaches and tailor rehabilitation strategies to optimize patient outcomes by considering different infarct sizes.

## Acknowledgment

We are highly thankful to Bethune First Hospital of Jilin University, Professor Ms. Wang, and Professor Ms.Ma for guiding us to finalize this article.Thanks to Xingqi Su, Lingmin Zhao,Jiulin You,LiangShu Feng,LiChong Zhu,Ying Chen,YuLei Hao,XinYu Wang, JiaChun Feng,the above authors for their joint efforts.

## Funding

None.

## Data availability

The data that support the findings of this study are available from the corresponding author upon reasonable request.

## ethics statements

The Ethics Committee of the First Hospital of Jilin University reviewed and approved the study, ensuring that it adhered to ethical principles and guidelines for human subject research. The approval number AF-IRB-032-02 indicates that the study has undergone thorough ethical scrutiny and met the necessary requirements.

The participants were provided with detailed information about the study, including its purpose, procedures, potential risks and benefits, confidentiality measures, and their rights as participants. They were given ample time to ask questions and clarify any concerns before voluntarily signing the consent form.

The written informed consent form documented the participant’s agreement to participate in the study and their understanding of the information provided. It also stated that they could withdraw from the study at any time without penalty.

To ensure confidentiality, the consent forms were securely stored and accessible only to the research team. Any identifying information was anonymized or coded to protect the participants’ privacy.The research team will continue to prioritize the welfare and rights of the participants throughout the study, maintaining ethical standards and promptly addressing any concerns that may arise.

Atients with acute ischemic stroke that presented within 72 h of onset from October 2017 to October 2019 were enrolled.

---

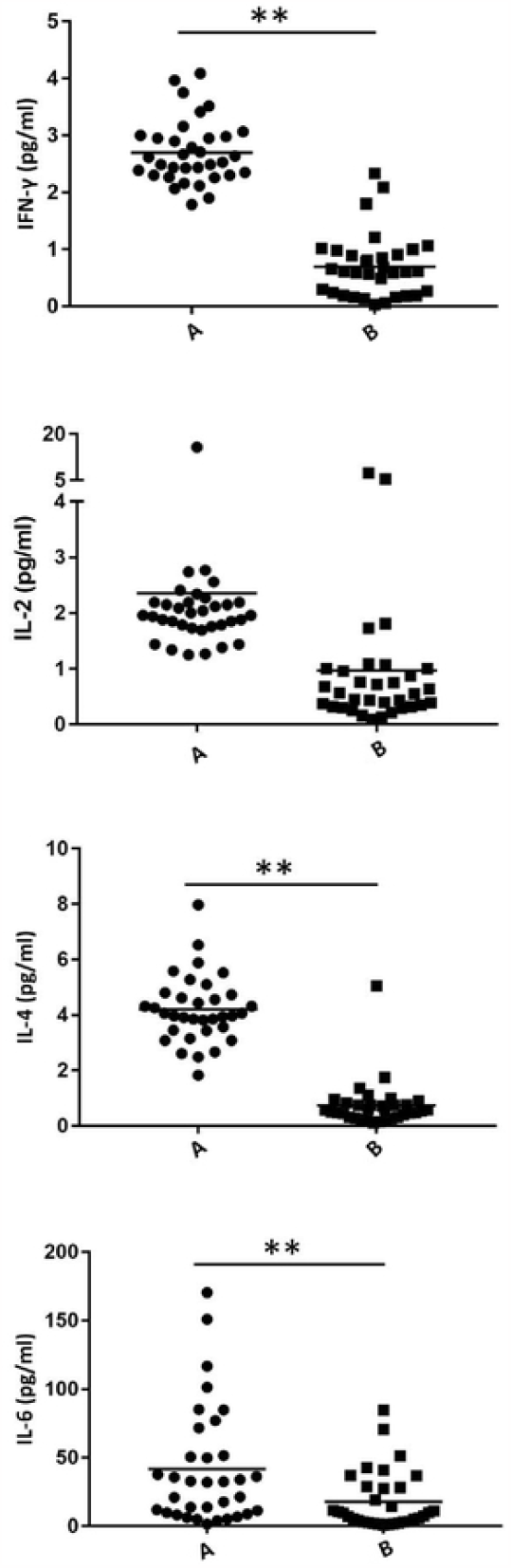

## Notes

### Competing Interest Statement

The authors have declared no competing interest.

### Clinical Trial

NCT03335111

### Funding Statement

The author(s) received no specific funding for this work.

### Author Declarations

The Ethics Committee of the First Hospital of Jilin University reviewed and approved the study

## References

1. Pan J, Li X, Peng Y. Remote ischemic conditioning for acute ischemic stroke: dawn in the darkness. Reviews in the Neurosciences. 2016;27.

2. Fernandez DM, Rahman AH, Fernandez NF, Chudnovskiy A, Giannarelli C. Single-cell immune landscape of human atherosclerotic plaques. Nature Medicine. 2019;25(10).

3. Pakzad B, Rajae E, Shahrabi S, Mansournezhad S, Saki N. T-Cell Molecular Modulation Responses in Atherosclerosis Anergy. Laboratory Medicine. 2020;(16).

4. Yi X, Zhu L, Sui G, Li J, Bao S. Inflammation and Endothelial Function Relevant Genetic Polymorphisms and Carotid Plaque in Chinese Population. Journal of Atherosclerosis and Thrombosis. 2020;27(9).

5. Korn T, Kallies A. T cell responses in the central nervous system. Nature Reviews Immunology. 2017;17(3):179–94. doi: 10.1038/nri.2016.144.

6. Becker KJ, Kalil AJ, Tanzi P, Zierath DK, Savos AV, Gee JM, et al. Autoimmune responses to the brain after stroke are associated with worse outcome. Stroke. 2011;42(10):2763–9. Epub 20110728. doi: 10.1161/strokeaha.111.619593.

7. Ren H, Liu X, Wang L, Gao Y. Lymphocyte-to-Monocyte Ratio: A Novel Predictor of the Prognosis of Acute Ischemic Stroke. J Stroke Cerebrovasc Dis. 2017;26(11):2595–602.Epub20170826.doi:10.1016/j.jstrokecerebrovasdis.2017.06.019.

8. Nam KW, Kim TJ, Lee JS, Kwon HM, Lee YS, Ko SB, et al. High Neutrophil-to-Lymphocyte Ratio Predicts Stroke-Associated Pneumonia. Stroke. 2018;49(8):1886–92. doi: 10.1161/strokeaha.118.021228.

9. Wang Y, Zhang JH, Sheng J, Shao A. Immunoreactive Cells After Cerebral Ischemia. Front Immunol.2019;10:2781.Epub 20191126. doi:10.3389/fimmu.2019.02781.

10. Jian Z, Liu R, Zhu X, Smerin D, Zhong Y, Gu L, et al. The Involvement and Therapy Target of Immune Cells After Ischemic Stroke. Front Immunol. 2019;10:2167. Epub 20190911. doi: 10.3389/fimmu.2019.02167.

11. Saino O, Taguchi A, Nakagomi T, Nakano-Doi A, Kashiwamura S, Doe N, et al. Immunodeficiency reduces neural stem/progenitor cell apoptosis and enhances neurogenesis in the cerebral cortex after stroke. J Neurosci Res. 2010;88(11):2385–97. doi: 10.1002/jnr.22410.

12. Butovsky O, Ziv Y, Schwartz A, Landa G, Talpalar AE, Pluchino S, et al. Microglia activated by IL-4 or IFN-gamma differentially induce neurogenesis and oligodendrogenesis from adult stem/progenitor cells. Mol Cell Neurosci. 2006;31(1):149–60. Epub 20051116. doi: 10.1016/j.mcn.2005.10.006.

13. Albini A, Marchisone C, Del Grosso F, Benelli R, Masiello L, Tacchetti C, et al. Inhibition of angiogenesis and vascular tumor growth by interferon-producing cells: A gene therapy approach. Am J Pathol.2000;156(4):1381–93.doi: 10.1016/s0002-9440(10)65007-9.

14. Wang S, Zhang H, Xu Y. Crosstalk between microglia and T cells contributes to brain damage and recovery after ischemic stroke. Neurol Res. 2016;38(6):495–503. Epub 20160531. doi: 10.1080/01616412.2016.1188473.

15. Cramer JV, Benakis C, Liesz A. T cells in the post-ischemic brain: Troopers or paramedics? J Neuroimmunol. 2019;326:33–7. Epub 20181113. doi: 10.1016/j.jneuroim.2018.11.006.

16. Jickling GC, Liu D, Ander BP, Stamova B, Zhan X, Sharp FR. Targeting neutrophils in ischemic stroke: translational insights from experimental studies. J Cereb Blood Flow Metab. 2015;35(6):888–901. Epub 20150325. doi: 10.1038/jcbfm.2015.45.

17. Maestroni GJ. Sympathetic nervous system influence on the innate immune response. Ann N Y Acad Sci. 2006;1069:195–207. doi: 10.1196/annals.1351.017.

18. Yuan M, Han B, Xia Y, Liu Y, Wang C, Zhang C. Augmentation of peripheral lymphocyte-derived cholinergic activity in patients with acute ischemic stroke. BMC Neurol. 2019;19(1):236. Epub 20191015. doi: 10.1186/s12883-019-1481-5.

19. Prass K, Meisel C, Höflich C, Braun J, Halle E, Wolf T, et al. Stroke-induced immunodeficiency promotes spontaneous bacterial infections and is mediated by sympathetic activation reversal by poststroke T helper cell type 1-like immunostimulation. J Exp Med. 2003;198(5):725–36. Epub 20030825. doi: 10.1084/jem.20021098.

20. Sekiya T, Yoshimura A. In Vitro Th Differentiation Protocol. Methods Mol Biol. 2016;1344:183–91. doi: 10.1007/978-1-4939-2966-5_10.

21. Shapouri-Moghaddam A, Mohammadian S, Vazini H, Taghadosi M, Esmaeili SA, Mardani F, et al. Macrophage plasticity, polarization, and function in health and disease. J Cell Physiol. 2018;233(9):6425–40. Epub 20180301. doi: 10.1002/jcp.26429.

22. Tschoe C, Bushnell CD, Duncan PW, Alexander-Miller MA, Wolfe SQ. Neuroinflammation after Intracerebral Hemorrhage and Potential Therapeutic Targets. J Stroke. 2020;22(1):29–46. Epub 20200131. doi: 10.5853/jos.2019.02236.

